# The Impact of National Context on COVID-19 Vaccine Hesitancy Across Europe: A Multi-Level Approach

**DOI:** 10.1101/2025.08.11.25333435

**Authors:** Michael Bergmann, Arne Bethmann, Tessa-Virginia Hannemann, Alexander Schumacher, Nikolaos Theodoropoulos

**Affiliations:** SHARE Survey Methodology, SHARE Berlin Institute, Germany; Hochschule für Technik und Wirtschaft des Saarlandes (htw saar), Germany; SHARE Germany, SHARE Berlin Institute; Department of Economics, University of Cyprus

**Keywords:** COVID-19, SHARE, vaccine hesitancy, multi-level, weighting, national context, macro indicators, 3C model, WHO, complacency, convenience, confidence

## Abstract

This study investigates vaccine hesitancy among the 50+ population in Europe and Israel during the COVID-19 pandemic, focusing on the role of national contexts alongside individual determinants. Utilizing data from the SHARE Corona Surveys (SCS1 and SCS2), the analysis includes over 48,000 respondents across 28 countries. The framework is guided by the WHO’s Complacency, Convenience, and Confidence (3Cs) model to explore factors influencing vaccination intent. Complacency examines perceived risks and visibility of the virus’s consequences, convenience evaluates accessibility and affordability of vaccines, and confidence assesses trust in vaccines, healthcare systems, and policymakers.

Key findings suggest that higher COVID-19 mortality rates and stricter containment measures reduce complacency and increase vaccine uptake. Greater healthcare expenditure and Human Development Index (HDI) scores enhance convenience, leading to higher vaccination rates. Additionally, trust in governments and health authorities as well as in the vaccine itself predict confidence, shaping vaccination intent.

The study reveals significant national disparities in vaccination attitudes and behaviours, linked to socio-economic factors and healthcare quality. By incorporating multivariate logistic regression models, the research highlights the interplay between individual and national factors, suggesting that successful vaccination campaigns require a holistic approach addressing both personal hesitations and systemic barriers. This research hence underscores the importance of public trust, robust healthcare systems, and targeted communication strategies to mitigate vaccine hesitancy and improve pandemic response outcomes.

## 1 INTRODUCTION

The rollout of vaccines against the novel coronavirus (COVID-19) marked a turning point in the pandemic (WHO, 2024). Vaccines were instrumental in saving lives and curbing the spread of the virus, allowing the return to “normal” life. Despite widespread availability, vaccine hesitancy persisted in several European countries. Older adults, prioritized for immunization due to their heightened risk of severe COVID-19 outcomes (WHO, 2023), were a particularly important group in this context, and are the focus of the present study. Bergmann et al. (2022) showed in a previous study, conducted on the same population, that vaccine hesitancy among the 50+ population in Europe was more prevalent among women and those with lower education levels. Living in rural areas, financial hardship, risk of poverty, as well as unemployment were further linked to a reluctance to vaccinate. Notably, vaccination rates varied dramatically across Europe, ranging from 25% to 98%, with the lowest rates in Eastern Europe and the highest in Southern Europe, though this pattern was not purely geographical (Jolly et al., 2025). This variation underscores the complex interplay of psychosocial, economic, and institutional factors that shape vaccine acceptance and that extend beyond individual characteristics.

This paper aims to highlight the importance of examining contextual variables like macroeconomic indicators and cultural influences alongside individual-level factors when analysing vaccine uptake probabilities globally. The analysis uses data from the first (SCS1) and the second SHARE Corona Survey (SCS2), conducted in 27 European countries and Israel in the summer of 2020 and 2021. The Complacency, Convenience, and Confidence (“3Cs”) model (WHO, 2014), which has previously provided a useful framework for examining national-level influences on vaccination uptake, serves as the theoretical basis for these analyses.

### Complacency

According to the Strategic Advisory Group of Experts (SAGE) Working Group on Vaccine Hesitancy, vaccine complacency can be thought of as the perceived risk of a vaccine-preventable disease or a severe course of the disease, i.e. if complacency is high, the perceived risk is low. Consequently, vaccination may not be regarded as an absolute necessity when it is weighed against the cost or effort. In the context of the COVID-19 pandemic, national complacency barriers to vaccination uptake are based on how respondents perceive the risk of a SARS-CoV-2 infection and of possible harm by the disease against how effectively they think the vaccine can protect them from an infection or at least serious symptoms. In this respect, the national death toll reflects the most severe consequences of the pandemic that often dominated national and international headlines. High death tolls also increase the likelihood of having personal experience with bereavement due to COVID-19, which in turn could alter risk perceptions (Cipolletta et al., 2022). Varbanova et al. (2024) found that higher vaccination rates were associated with more stringent public health measures. Therefore, a positive relationship between higher death rates per COVID-19 cases in a country as well as more severe measures taken to contain infection rates by country leaders can be assumed.

### Convenience

Convenience is defined by the extent to which the physical ability to get vaccinated, the affordability and willingness to pay, the geographical accessibility of vaccines, the ability to understand the purpose of vaccination, and the appropriateness of the quality of vaccination services, as well as the time, place, and cultural context, affects the decision to be vaccinated. General socio-economic context influences on convenience can be regarded through several pathways. Firstly, capacity to roll out a vaccine and to campaign for its adoption is expected to be higher in well-off nations with well-functioning health systems. As for the COVID-19 pandemic, there was a common initiative by the EU commission to buy and distribute vaccines to member states. In this respect, differences in affordability across countries can be seen as a minor issue in Europe, although the quality and the responsiveness of vaccination services within the cultural context differed a lot across different regions (Jolly et al., 2025). Multiple macro-level studies have explored these contextual determinants. Rughiniş et al. (2022) found that human development, particularly life expectancy, as well as trust in healthcare professionals were strong predictors of national vaccination rates. In contrast, economic indicators such as the gross domestic product (GDP) and education alone had limited predictive power. A study by Lamot and Kirbiš (2024) also found that economic development (measured by GDP) and income equality (lower Gini coefficient) moderated the determinants of vaccination, in that a higher GDP and lower Gini coefficient lead to more successful vaccination outcomes. Further, Roghani (2021) demonstrated that while GDP was strongly correlated with early vaccine distribution, the Human Development Index (HDI) showed a weaker association. Pronkina and Rees (2022) added nuance by identifying lower vaccination rates among those who are financially risk-averse, frequent in religious practice, or low in interpersonal trust—suggesting that values and social capital are influential. Additionally, lower vaccination rates in former communist countries were linked to diminished social capital (Pronkina et al., 2023). In the context of convenience of access to vaccination, we thus can assume that high investment in the health care system will in general improve services. It might also facilitate more and closer contact among the population with health care practitioners and facilities. Both factors should improve trust in the health care system and offer more opportunities for health-care professionals to address citizens about vaccines. We would thus expect a positive effect of country-level health expenditures on the convenience of receiving a vaccination and therefore higher commitment to vaccinate in these countries.

### Confidence

Finally, confidence is defined as trust in three key areas: the effectiveness and safety of vaccines, (2) the system that delivers them, including the reliability and competence of the health services and health professionals, and (3) the motivations of the policymakers who decide on the needed vaccines. For a vaccination campaign it is therefore crucial that individuals have positive attitudes towards the safety and effectiveness of the vaccines, as well as the political actors and health authorities. They need to, for example, be satisfied with governmental policy actions related to handling the pandemic, such as providing them with all necessary information on the vaccination. Low confidence in government response and distrust in vaccine safety were key predictors of hesitancy in Portugal (Gomes et al., 2022). Similarly, Franic (2022) found that dissatisfaction with democratic institutions and low trust in state bodies were significant correlates of vaccine hesitancy across Europe. Political ideology also plays a role. Borga et al. (2022) and Bade et al. (2024) observed that lower trust in government and greater support for populist or right-wing parties was associated with higher vaccine hesitancy. Further, Lamot and Kirbiš (2024) analysed data from twenty-six countries and found that the interaction of healthcare satisfaction, trust in institutions, and conspiracy beliefs with national-level indicators significantly affected vaccination rates. Satisfaction with healthcare had a stronger positive effect in less corrupt, more individualistic countries, while conspiracy beliefs had stronger negative effects in less developed and more collectivist societies. We therefore expect higher levels of trust and satisfaction with the handling of the pandemic to lead to lower rates of vaccine hesitancy.

Considerations of the 3Cs theoretical framework and the existing literature have led to the following hypotheses:

1. (a) Due to the visibility of the consequences of the virus, a higher death rate related to COVID-19 should result in lower complacency and thus higher intention to vaccinate. (b) Similarly, stricter measures to contain the spread of the virus, which increase the population’s sense of affectedness, should decrease complacency and increase the intention to vaccinate.
2. (a) Greater health expenditure can lead to greater access to health care, which should increase the convenience of the vaccine. We would therefore expect a higher percentage of GDP spent on health care to result in higher intention to vaccinate. (b) Similarly, a better standing in the HDI should result in greater national access to healthcare and higher literacy, increasing convenience as well as intention to vaccinate.
3. (a) Confidence in the vaccine’s safety and effectiveness must be considered a pre-requisite to get vaccinated which why is why we assume that higher levels of confidence are related to higher levels of vaccination intent. (b) As national governments, health authorities and similar institutions need to communicate and promote vaccination policies, it is crucial that the respective population has confidence in their handling of the pandemic. Higher confidence in these institutions should therefore lead to more widespread adoption of the vaccine.

## 2 DATA AND METHODS

### 2.1 Data Source

In the following analyses, we use released data from the first and the second SHARE Corona Survey (SHAREERIC, 2024h, 2024j) that were fielded, respectively, during summer 2020 and 2021 in 28 countries (Austria, Belgium, Bulgaria, Croatia, Cyprus, Czech Republic, Denmark, Estonia, Finland, France, Germany, Greece, Hungary, Israel, Italy, Latvia, Lithuania, Luxembourg, Malta, Netherlands, Poland, Portugal, Romania, Slovakia, Slovenia, Spain, Sweden and Switzerland). The SHARE Corona Survey is a Computer Assisted Telephone Interview (CATI) created in reaction to the COVID-19 crisis. It collected data on the living situation of people aged 50 years and over across Europe and Israel during the pandemic (Bergmann et al., 2024; Scherpenzeel et al., 2020). The gross sample of the first SHARE Corona Survey consisted of all panel respondents who had been eligible for the regular SHARE Wave 8, while the second SHARE Corona Survey reinterviewed these respondents to enable the examination of changes between the start of the pandemic and the situation about one and a half years later. Verbal consent, emphasizing the voluntary nature of participation and the confidentiality of data (e.g. researchers have no access to information that could identify individual participants during or after data collection), was obtained from all participants.

Respondent information from the regular SHARE panel study (for a general overview, see Börsch-Supan et al., 2013) was added during our analyses to provide long-term information on stable respondent characteristics (SHARE-ERIC, 2024a, 2024b, 2024c, 2024d, 2024e, 2024f, 2024g, 2024i). The average response rate based on eligible respondents who participated in the first SHARE Corona Survey was 79%. In the second SHARE Corona Survey, an average retention rate (excl. recovery of respondents) of 86% was achieved. To avoid selectivity, our analyses are based on 48,058 respondents aged 50 years and older who participated in both SHARE Corona Surveys.

### 2.2 Measures

We operationalized vaccination status and intent to get vaccinated based on two consecutive questions: First, respondents were asked whether they had been vaccinated against COVID-19 at least once. Second, of those who had not yet been vaccinated, information on their intention to do so was requested inquiring whether they already had scheduled an appointment for vaccination, wanted to get vaccinated, did not want to get vaccinated, or were still undecided. Respondents who answered “Don’t know” in the question on vaccination intent were categorized as undecided. Our main dependent binary variable then distinguishes those who are vaccinated or want to be, from those who are undecided or do not want to get the vaccine.

To properly analyse the differences in vaccine uptake across Europe, we examine the impact of both individual- and country-level indicators. Regarding individual characteristics, we build on previous research (Bergmann & Wagner, 2021; Bergmann et al., 2022) and included three domains of influential factors, namely socio-demographics, physical and mental health indicators, and respondents’ living and economic conditions.

Regarding socio-demographics, we used the respondents’ sex (0: male, 1: female) and their age at interview. Further, we coded the level of education attained based on the Internal Standard Classification of Education 1997 (ISCED-97). Respondents were then grouped into three categories: primary education (ISCED-97 score: 0-2), secondary education (ISCED-97 score: 3), and post-secondary education (ISCED-97 score: 4-6).

As physical and mental health indicators, we considered respondents’ self-rated health (0: poor/fair, 1: good, 2: very good/excellent), whether they had at least one diagnosed illness, and if respondents were affected by mental health issues, such as feeling depressed, anxious, lonely, or having had trouble sleeping. To assess the extent to which respondents had been affected by COVID-19, we created a 3-point variable, accounting for “not affected” (no one affected close to the respondent), “mildly affected” (someone close to the respondent tested positive or developed symptoms for COVID-19), and “severely affected” (someone close to the respondent had been hospitalized or died due to COVID-19). Concerning living and economic conditions, we used information on the respondents’ type of living area (0: rural area, 1: urban area like a large town or big city). Furthermore, we measured respondents’ subjective economic situation by a question that asked the degree to which respondents can make ends meet (0: with great/some difficulty, 1: fairly easily/easily) and their objective economic status by measuring respondents’ risk of poverty (disposable income is less than the equivalence of 60% of the national median). Finally, we included a measure related to whether the respondent was employed (including self-employment), retired or had another non-working status (unemployed, permanently sick or disabled and homemaker) at the beginning of the outbreak of COVID-19.

Regarding macro-level indicators, we first used the ratio of cumulated deaths per 100,000 inhabitants in a country during the first wave of the COVID-19 pandemic, i.e. up to June 30, 2020, to measure complacency barriers to COVID-19 vaccine uptake, specifically including the role of media attention at the beginning of the pandemic. The number of deaths is taken from the Oxford COVID-19 Government Response Tracker (OxCGRT; see Hale et al., 2021). Also based on these data, we used the so-called stringency index to assess differences in national policy responses toward the pandemic. The index records the strictness of “lockdown style” policies, which primarily restrict people’s behaviour and in particular in-person contacts. It aggregates policy responses about school and workplace closings, cancelling of public events, restrictions on gatherings, closure of public transports, stay-at-home requirements, restrictions on internal movement, international travel controls and public information campaigns (Hale et al., 2021). The stringency index is the average of the aforementioned policy indicators over the recorded days. It ranges from 0 to 100, with greater values indicating greater strictness. We classified a country as under severe measures at a stringency of over 50 points and counted the number of days under severe measures before the vaccine became available. By this, we were able to match precisely the country-specific context information on the pandemic to the respondents’ behaviour.

To measure vaccine convenience, we used components of the Human Development Index (HDI)^1^, which captures average achievement in three key dimensions: a long and healthy life, being knowledgeable, and having a decent standard of living. The health dimension is assessed by life expectancy at birth; the education dimension is measured by expected years of schooling for children of school entering age; and the standard of living dimension is measured by the gross national income (GNI) per capita in constant 2017 purchasing power parity (PPP) terms. We additionally included current health expenditure as a percentage of the national gross domestic product (GDP) in 2021. This measure is taken from the WHO database^2^ and provides an indication of the level of resources channelled to health relative to other uses.

Finally, to measure vaccine confidence, we used attitudes on vaccination against COVID-19 from the Flash Eurobarometer 494 that was collected in May 2021^3^. More precisely, we use answers from respondents 50+ on the safety and effectiveness of vaccines as well as on their trust in the national government and in health authorities to give reliable information on COVID-19 vaccines. Additionally, we used the Corruption Perceptions Index (CPI)^4^, which ranks countries by perceived public sector corruption on a scale from 0 (highly corrupt) to 100 (very clean), as an indicator of distrust in government and its handling of the pandemic, a factor that can drive vaccine hesitancy.

### 2.3 Statistical Analyses

To address our research questions outlined above, we first looked descriptively at the macro-level variables to better understand their bivariate association with respondents’ vaccination intention aggregated at the country level. To further analyse the effect of country-level variables using the “3Cs” indicators (complacency, convenience and confidence) on the respondents’ intention to get vaccinated, we included the following individual-level control variables to our multilevel logistic model: socio-demographic variables (age, sex, level of education), physical and mental indicators (self-rated health, diagnosed illnesses, mental health strains, direct affectedness), and respondents’ living and economic conditions (area type, subjective and objective economic situation, working status). The multilevel approach enables analysing variables from different levels simultaneously by properly considering the statistical dependencies between the observations to adjust standard errors, which are likely to be biased if the hierarchical structure of the data is ignored (Hox et al., 2010). The dependent variable, vaccination intent, was treated as binary in the multilevel model, with the customary logit function defined as *logit*(*x*) = *ln*[*x*/(1–*x*)]. The predicted value for *P*_*ij*_ in the general logistic multi-level model was extended to include explanatory variables *X* at the individual level and country-level variables *Z*. It can be written as follows:

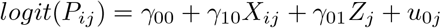

where the random intercept *γ*_00_ is shared by all countries, while the residual term *u*_0*j*_ is specific to country *j* and assumed to follow a normal distribution with variance 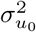 To quantify the extent to which vaccine intent varies between countries, the intraclass correlation coefficient (ICC) was calculated as follows in the intercept-only model without explanatory variables:

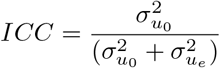

where 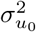 is defined as the country variance at level two, and the individual variance at level one, 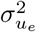, was fixed to 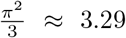 in logistic multilevel regressions (Goldstein et al., 2002; Hox et al., 2010). Higher values of the ICC indicate a stronger influence of country differences on vaccine uptake. Because variance components in multilevel logistic regressions cannot be directly compared across models with and without explanatory variables due to the fixed level-1 variance, we followed the approach by Hox et al. (2010) and calculated a scale correction factor for each model with explanatory variables. With this correction, we were able to assess the amount of variance explained separately at each level.

As we are primarily interested in the effect of macrolevel predictors, we applied a two-level logistic regression model with random intercepts and fixed slopes. Average marginal effects (AME) were used to facilitate comparisons of the indicators in the multivariate model. Analyses were conducted using Stata 17 and the melogit command, which is based on a maximum likelihood estimation procedure using adaptive quadrature with seven integration points. Results are based on robust standard errors and using the provided calibration weights at the individual level. In this respect, we normalised the individual-level survey weight to the cluster sample size (i.e. the weights have a mean of 0 and a standard deviation of 1 in each country) and trimmed a few very large weights (on average, 13 cases per country; n=331) to the 99^th^ percentile, in order to avoid huge standard errors and thus enable meaningful conclusions to be drawn. Further, we compiled country-level weights based on the total 50+ population in the participating countries. Following Asparouhov (2006) and Carle (2009), we rescaled the country populations relative to the total sample sizes in the data to give more (less) weight to countries, which are under-(over-)represented in the data. By this, we can draw proper conclusions on the 50+ population in Europe, considering nonresponse at the individual level and the relative size of the participating countries on the country level in our estimations.

## 3 RESULTS

### 3.1 Bivariate Analysis

We start our analysis with the bivariate correlations depicted in Figure 1 between the vaccination rate and the context measures we constructed to represent the parts of the 3C model.

**Figure 1:**
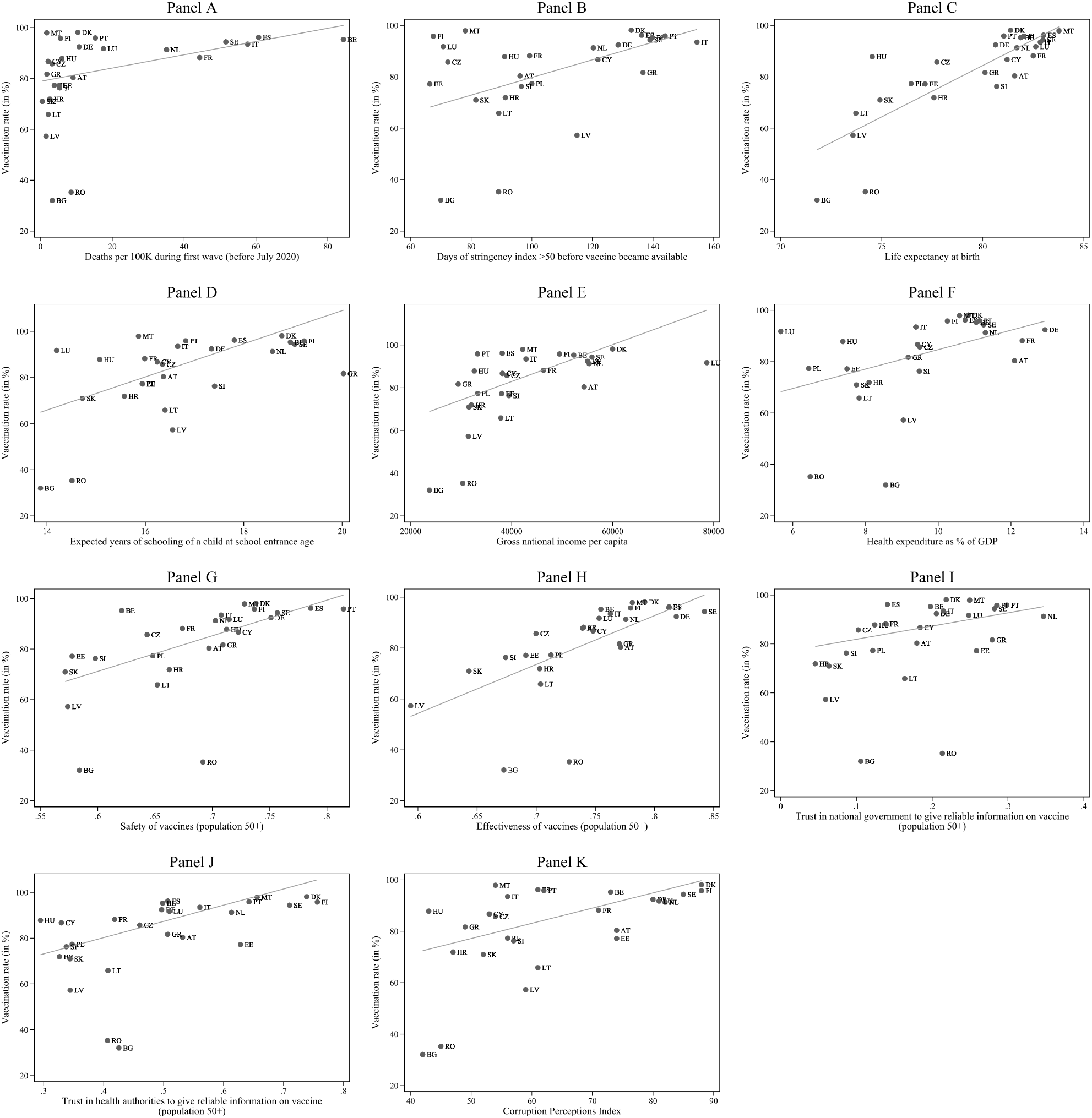
Bivariate correlations between macro indicators and vaccination rate, country level Data: SHARE Wave 9 COVID-19 Survey 2, Release 9.0.0 (n=45,929, weighted).

#### COVID-19-related context (Complacency)

We argued that national death tolls drive COVID-19 risk perception both through heightened media attention and increased likelihood of personal experience with bereavement. Therefore, we used national death tolls as an indicator counteracting complacency and expected the number of deaths during the first wave of the pandemic to be connected to vaccination acceptance. Figure 1 shows a positive correlation between the number of casualties per 100,000 inhabitants and the rate of vaccination intent at the country-level (r=.44, see Figure 1, Panel A). This correlation with early COVID-19-related deaths represents just under a fifth of the variance in vaccination rates (R^2^=.20). Excluding the two outliers Romania and Bulgaria increases the correlation slightly (r=.54, R^2^=.29). This seems to indicate that COVID-19 related deaths during the first months of the pandemic played a role in the decision to vaccinate. This points to media attention being a relevant mechanism, although actual death tolls overall were still comparatively low at that time and being affected personally by a death in the closer social network due to COVID-19 was quite unlikely.

As a further argument for complacency driving vaccination hesitancy, we proposed that containment measures would signal a greater risk arising from COVID-19 infections and would thus increase vaccination uptake. We used the number of days with a stringency index greater than 50 (out of 100 points) until the vaccine became available as a measure. We would expect that vaccination acceptance rates rise with the number of days spent under severe measures. Indeed, we observe a positive correlation of stringency and vaccination rates of r=.59 (see Figure 1, Panel B). This is stronger than the association with the COVID-19 death toll and explains more than a third of the variance in vaccination acceptance rates, underlining the relevance of lockdown measures with regards to complacency toward vaccination. Stringency seems to be more closely related to vaccination acceptance rates than the death toll of the pandemic, but both variables are equally correlated to each other as to vaccination acceptance.

#### Socio-economic context (Convenience)

The socio-economic context determines both state capacity and the quality of healthcare services. Consequently, these factors influence vaccine rollout efforts as well as contact to healthcare and thus increase the ease of getting vaccinated. We therefore used the subscales of the Human Development Index (HDI) as well as health expenditures as indicators of vaccination convenience and expected a positive association of vaccine acceptance with each of them.

The HDI consists of the three sub-indicators life expectancy at birth, expected years of schooling of a child at school entrance age, and gross national income per capita. The greatest factor driving vaccination rates is life expectancy at birth, with a correlation of r=.81 (see Figure 1, Panel C). In other terms, life expectancy could explain almost two thirds of the country variance in vaccination acceptance rates. Years of schooling is also significantly related to vaccination outcomes with r=.62 (see Figure 1, Panel D). GNI per capita seems to play a relatively minor role, with a correlation of r=.56 (see Figure 1, Panel E).

As investment in health infrastructure should enhance a country’s ability to roll out the vaccine and inform the public about it, we expect a positive correlation of health expenditure as a share of the national gross domestic product (GDP) and vaccination acceptance rates. Our findings show that the association between these two variables is indeed positive, as indicated by the correlation coefficient of r=.56 (see Figure 1, Panel F). Health expenditure thus can explain a little over a third of the variance between countries.

#### Attitudes towards the vaccine and relevant authorities (Confidence)

Finally, the confidence component of the WHO concept was examined. More specifically, we analysed the relationship between vaccination hesitancy and each of the measures in the area described in section 2.2: perceived safety and efficacy of the vaccine, trust in the government and in the health authorities, as well as the perceived level of corruption in the respective country.

If we consider the full set of countries, we see a medium correlation of r=.52 between the perceived safety of vaccines and the vaccination rate (see Figure 1, Panel G). Once more, Bulgaria and Romania must be considered outliers due to their very low vaccination rates. Excluding them leads to a higher correlation (r=.71), indicating that the perceived safety of the vaccine is positively related to individuals’ motivation to get vaccinated as they perceive the benefits to outweigh the risks. Apart from being safe, i.e. not causing harm, a vaccine must also be expected to be effective in preventing an actual infection for it to be accepted. This is indeed what Figure 1 (Panel H) suggests, depicting the relationship between the perceived effectiveness of the COVID-19 vaccine and the vaccination acceptance rate. With a correlation of .61 for all countries and .77 when excluding Romania and Bulgaria this indicator for the confidence area shows a slightly stronger relationship to vaccination acceptance than the perceived safety of the vaccine.

In addition to confidence in the vaccine itself, we looked at the confidence people have in the institutions in charge of the political handling of the pandemic, as well as the practical distribution of the vaccine, which we measure through trust in the government and in the health authorities, respectively. If we look at the bivariate relationship between the vaccination acceptance rate and trust in national government/health authorities regarding available information on vaccines, we see a positive correlation for both indicators (r=.23 for trust in government, see Panel I, and r=.47 for trust in health authorities, see Panel J). Countries with a higher average level of trust, e.g. Netherlands or Denmark and Finland, tend to be in the top group regarding vaccination rates. Instead, Eastern European countries, such as Croatia, Latvia or Slovakia, show lower trust levels and exhibit comparatively lower vaccination rates. Romania and Bulgaria have by far the lowest vaccination rates, although they are not among the countries with the lowest trust levels.

Assuming that a higher level of corruption will likely come with a reduction in confidence in political institutions and their ability to handle the pandemic, we lastly looked at the Corruption Perception Index (CPI) with higher values indicating less perceived corruption. Figure 1 (Panel K) shows a correlation of .52 between the CPI and vaccination intent. This goes down to .36 when Bulgaria and Romania are excluded. Overall, however, it suggests that vaccination acceptance is higher in countries with lower levels of (perceived) corruption.

### 3.2 Multivariate Analysis

To paint a complete picture of the influencing factors on our respondents’ willingness to be vaccinated, we computed a multilevel hierarchical model combining individual characteristics with context effects. All models are compared to a null model, which only takes into account country differences. We then assessed the amount of variance explained by the additional explanatory variables, which were added stepwise: In model 1, we assessed the coefficients of individual characteristics, i.e. age, gender, education, physical and mental health, being affected by COVID-19, urban or rural area of living, ability to make ends meet, poverty risk and working status. In models 2 through 4, we sequentially added the country-level variables, representing complacency, convenience and confidence, to see how the context effects hold up against individual-level controls. Finally, we calculated a full model (model 5), combining individual-level predictors with all country-level indicators (the complete models with all parameter estimates can be found in Table A1 in the Appendix). To avoid multicollinearity, we standardized the country-level variables and compiled additive indices representing the three different dimensions of complacency, convenience, and confidence for the final model. In this respect, we divided the confidence dimension into one reflecting confidence in the vaccine and another one reflecting confidence in authorities (including the corruption perceptions index).

Table 1 summarizes the estimations of the different models, i.e. the null model, the random intercept model with individual (level-1) predictors and the random intercept model with all country (level-2) predictors. It shows that vaccination intent differed significantly between countries. This is reflected in the ICC of the intercept-only model, which was 1.18 / (3.29 + 1.18) = 26.5%, that is, about one fourth of the total variance in vaccination intent was attributable to differences between countries. The intercept-only model also gives us a benchmark value of the deviance (i.e., the degree of misfit of the model), which can be used to compare models with additional explanatory variables. From Table 1 it can be concluded that the deviance goes down when including explanatory variables at the different levels, thus indicating an improved model fit. A formal chi-square test to evaluate the difference of the deviances indicated significant improvements of the model fit when including all level-1 and level-2 predictors, respectively (from 5153.60 in the intercept-only model to 946.57 in the full model with all level-1 and level-2 predicators).

**Table 1:**
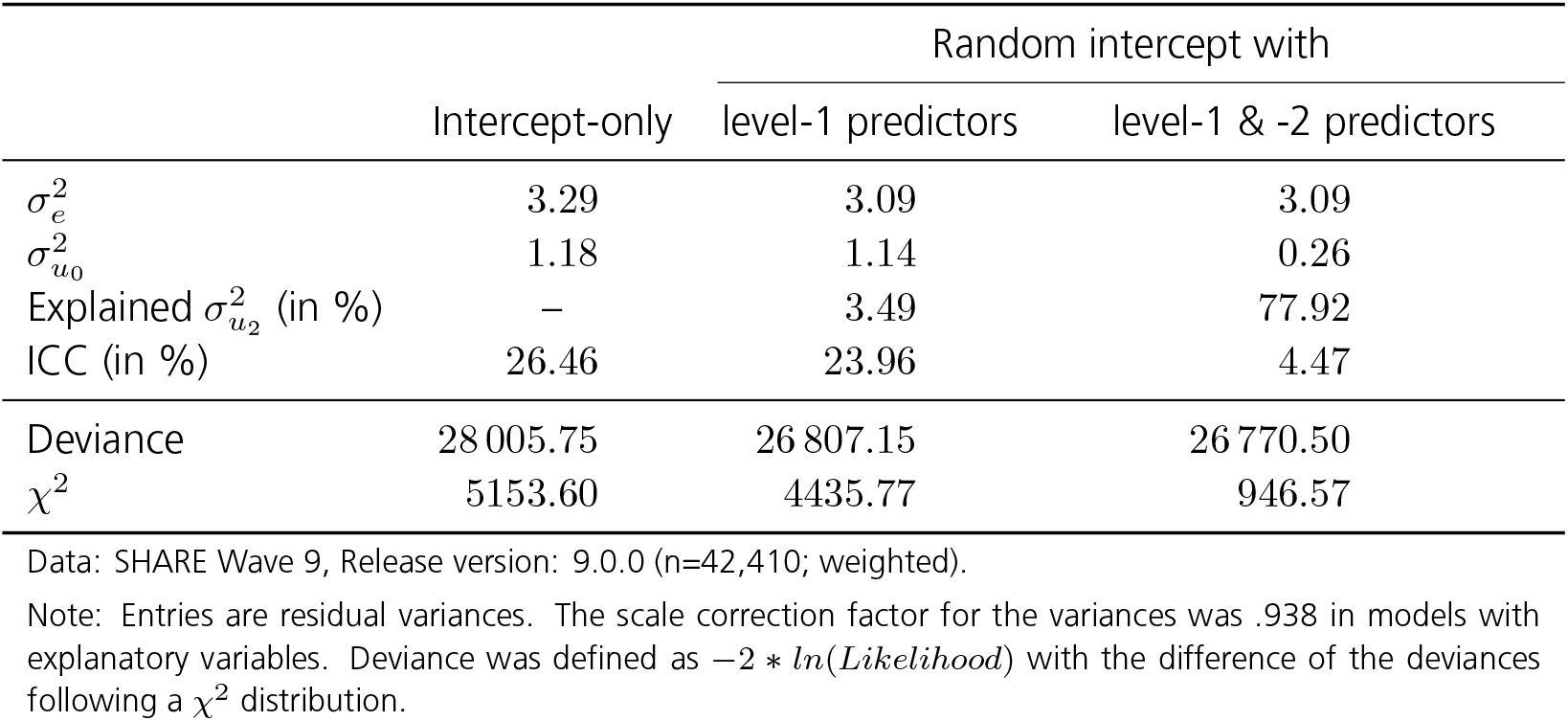
Rescaled estimates of individual (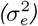) and country residual variance (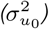) of sequential random intercept models regarding respondents’ answers on vaccine intent.

To further analyse how much residual error is left at the distinct levels and to assess the amount of explained variance at the different levels in multilevel logistic regressions, we needed to bring the sequential models to the same scale (Hox et al., 2010). Table 1 presents the rescaled variances from our multilevel logistic regression models. We see that after including respondent characteristics at the individual level and context characteristics at the country-level, the residual error variance at the country level„ decreased compared to the intercept-only model from 1.18 to .26. We can interpret the respective differences as the amount of variance explained by introducing explanatory variables at the different levels: The rescaled explained variance at the country level was about 3.5% after including individual characteristics and about 77.9% after including individual and country characteristics.

This result shows that the amount of variance explained by individual respondent characteristics at the country level was rather small, which reflects the fact that the included level-1 explanatory variables were distributed rather equally across countries. This encouraged us in our decision to assume fixed slopes across countries. Adding the country-level explanatory indices, representing the three dimensions of complacency, convenience and confidence, did not change the residual variance at the first level because the second-level variables cannot predict individual-level variation. However, the country-level residual variance went down to .26, which translates into nearly 78% of explained variance at the country level by both respondent and country predictors. Most of the predictive power of the model can hence be attributed to the included context predictors that differ across countries. This can also be seen by the clearly decreased ICC of less than 5% in the final model with all level-1 and level-2 predictors included.

#### Effects of the 3C indicators on vaccination intent

Figure 2 graphically presents the average marginal effects (AMEs) of the respondent- and country-level predictors for the multilevel logistic regression model. It shows that respondents are more likely to be vaccinated or want to be if they have a post-secondary level of education (as opposed to a primary level), report a diagnosed physical illness or have been severely affected by a COVID-19 infection of a close friend, family member or themselves. On the other hand, we see a significant lower vaccination intent among respondents having at least some difficulties in making ends meet, are at risk of poverty, and are (self-)employed or currently not working (incl. being unemployed, permanently sick/disabled or a homemaker).

**Figure 2:**
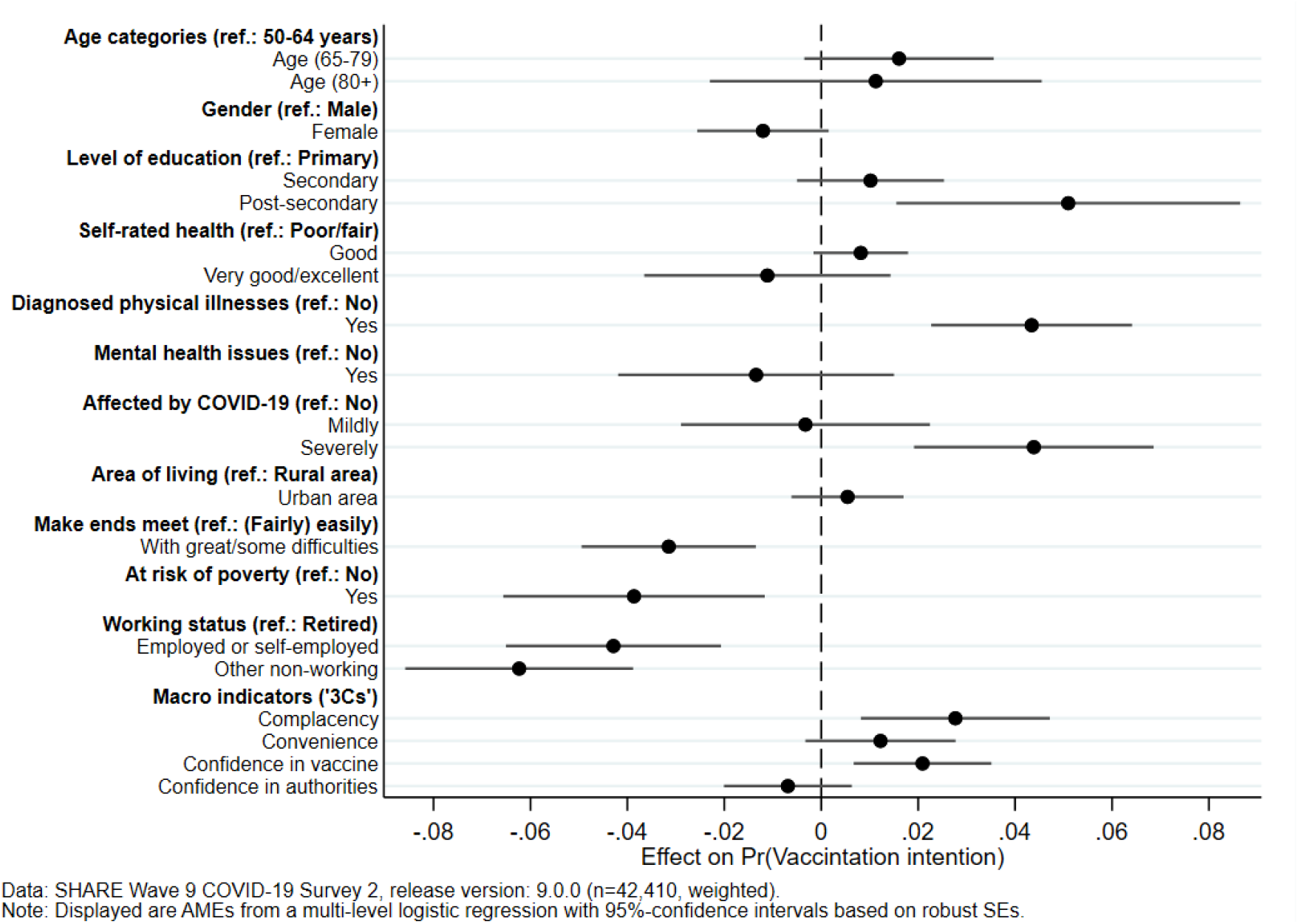
Multilevel model with individual characteristics and context effects

When turning to the country-level predictors, the dimensions of complacency and confidence in the vaccine show significant positive effects, indicating a higher probability of vaccination, whereas convenience is not significantly related to vaccination intent. Confidence in authorities is negatively (although not significantly) correlated with vaccination intent.

This generally underlines our expectation derived from the “3C” theory of vaccination motivation, proving its explanatory power for vaccination intent. Returning to the hypotheses presented in Section 1, we confirm Hypothesis 1: our complacency indicator, constructed from the number of deaths (per 100.000 inhabitants of a specific country during the first COVID-19 wave) and the severity of lockdown measures (number of days with elevated stringency before the vaccine became available), shows a statistically significant increase of about 2.8 percentage points in individual vaccination intent.

Hypothesis 2 postulates that the convenience of the vaccination should increase with a better funded health-care system (as proportion of national GDP) as well as a better national standing in the components of the HDI. While there is some positive correlation of the respective items with the intent to be vaccinated and the point estimate for the effect of the convenience index is positive also in the multivariate model, the latter does not reach the level of statistical significance. Therefore, our analyses do not provide statistical support for this hypothesis.

Lastly, we formulated the hypothesis that higher confidence should positively relate to vaccination intent. This is true and statistically significant for confidence in the vaccine itself (safety and effectiveness), but not for the confidence in authorities (measured through trust in governance and health authorities, as well as perceived corruption in the country), which shows a slightly negative, non-significant effect. We therefore find only partial support for our third hypothesis with an average increase in vaccination intent of about 2.1 percentage points for confidence in the vaccine.

## 4 CONCLUSION

During the COVID-19 pandemic, vaccines became critical tools in reducing mortality and protecting vulnerable populations. Despite widespread availability, vaccine hesitancy persisted, also among adults aged 50+ (see, e.g., Bergmann et al., 2022). This paper examines macro-level determinants of vaccination intent across Europe and Israel, using data from the SHARE Corona Surveys. We find that vaccine hesitancy among older adults during the COVID-19 pandemic was shaped not only by individual characteristics but also by macro-level contextual factors. While previous research has focused predominantly on personal attributes such as gender, age, education, and health status, our analysis reveals that national-level conditions account for most of the cross-country variation in vaccination intent. For instance, we find that only 3.5% of the cross-country variation in vaccination uptake can be explained by individual characteristics. In contrast, nearly 78% of the variation is explained by country-level context factors.

Applying the WHO’s 3Cs model (complacency, convenience, and confidence) in a multivariate setting suggests that complacency and convenience increased the probability of getting vaccinated. Specifically, stringent public health measures and high early death tolls were significantly associated with reduced complacency and greater acceptance of vaccines. Also, confidence in vaccine safety and effectiveness as opposed to confidence in authorities significantly increased the likelihood of vaccination.

Our findings underscore that successful vaccination strategies require a holistic, multi-level approach. Individual behaviour is embedded within broader institutional and cultural environments, and thus, public health campaigns must align clear, science-based messaging with robust healthcare delivery. In practical terms, this means that policymakers must go beyond targeting demographic subgroups with behavioural nudges and address structural factors such as healthcare access and risk communication. Especially for older populations, who are both more vulnerable and more dependent on public systems, sustained investments in health infrastructure and confidence in vaccine safety are vital.

Like other studies, our analysis has some limitations. Although we included important macro-level variables in line with the WHO 3Cs model, omitted variable bias may have been introduced by other contextual factors, such as the influence of social media or exposure to misinformation, which played an important role during the pandemic. Another limitation is that our analyses are based on respondents aged 50 years and over. This must be carefully considered when drawing generalized conclusions about the wider European population based on our findings. Future studies should therefore focus on younger age groups, particularly when analysing vaccinations other than those for COVID-19.

As older people have been affected most severely by this disease, our analysis can, however, serve as a starting point for future research in this area. By integrating micro- and macro-level insights, this study advances our understanding of vaccine hesitancy in Europe and contributes to the broader literature on public health preparedness. As future pandemics are likely, our results highlight the urgent need for resilient, equitable, and trust-based vaccination systems across diverse national contexts. Furthermore, the introduction of vaccinations against the coronavirus could also impact general attitudes towards vaccination (see, e.g., Lunz Trujillo et al., 2024), making it a vital component of a global strategy to enhance the vaccination status of vulnerable groups.

## Data Availability

The SHARE release data used in this paper are available at https://share-eric.eu/data/data-access.
The Oxford COVID-19 Government Response Tracker (OxCGRT) data are available at https://www.bsg.ox.ac.uk/covidtracker
The data for the Human Development Index (HDI) are available at https://hdr.undp.org/data-center/documentation-and-downloads
WHO database data are available at https://apps.who.int/nha/database/ViewData/Indicators/en
Data from Flash Eurobarometer 494 are available at https://europa.eu/eurobarometer/surveys/detail/2512
Data on the Corruption Perceptions Index (CPI) are available at https://www.transparency.org/en/cpi/2021

## ACKNOWLEDGEMENTS

This paper uses data from SHARE Waves 1, 2, 3, 4, 5, 6, 7, 8 and 9 (DOIs: 10.6103/SHARE.w1.900, 10.6103/SHARE.w2.900, 10.6103/SHARE.w3.900, 10.6103/SHARE.w4.900, 10.6103/SHARE.w5.900, 10.6103/SHARE.w6.900, 10.6103/SHARE.w6.DBS.100, 10.6103/SHARE.w7.900, 10.6103/SHARE.w8.900, 10.6103/SHARE.w8ca.900, 10.6103/SHARE.w9.900, 10.6103/SHARE.w9ca900, 10.6103/SHARE.HCAP1.100); see Börsch-Supan et al. (2013) for methodological details. The SHARE data collection has been funded by the European Commission, DG RTD through FP5 (QLK6-CT-2001-00360), FP6 (SHARE-I3: RII-CT-2006-062193, COMPARE: CIT5-CT-2005-028857, SHARELIFE: CIT4-CT-2006-028812), FP7 (SHARE-PREP: GA N°211909, SHARE-LEAP: GA N°227822, SHARE M4: GA N°261982, DASISH: GA N°283646) and Horizon 2020 (SHARE-DEV3: GA N°676536, SHARE-COHESION: GA N°870628, SERISS: GA N°654221, SSHOC: GA N°823782, SHARE-COVID19: GA N°101015924) and by DG Employment, Social Affairs & Inclusion through VS 2015/0195, VS 2016/0135, VS 2018/0285, VS 2019/0332, VS 2020/0313, SHARE-EUCOV: GA N°101052589 and EUCOVII: GA N°101102412. Additional funding from the German Federal Ministry of Education and Research (01UW1301, 01UW1801, 01UW2202), the Max Planck Society for the Advancement of Science, the U.S. National Institute on Aging (U01_AG09740-13S2, P01_AG005842, P01_AG08291, P30_AG12815, R21_AG025169, Y1-AG-4553-01, IAG_BSR06-11, OGHA_04-064, BSR12-04, R01_AG052527-02, R01_AG056329-02, R01_AG063944, HHSN271201300071C, RAG052527A) and from various national funding sources is gratefully acknowledged (see ww.shareeric.eu).

---

**Table A1:**
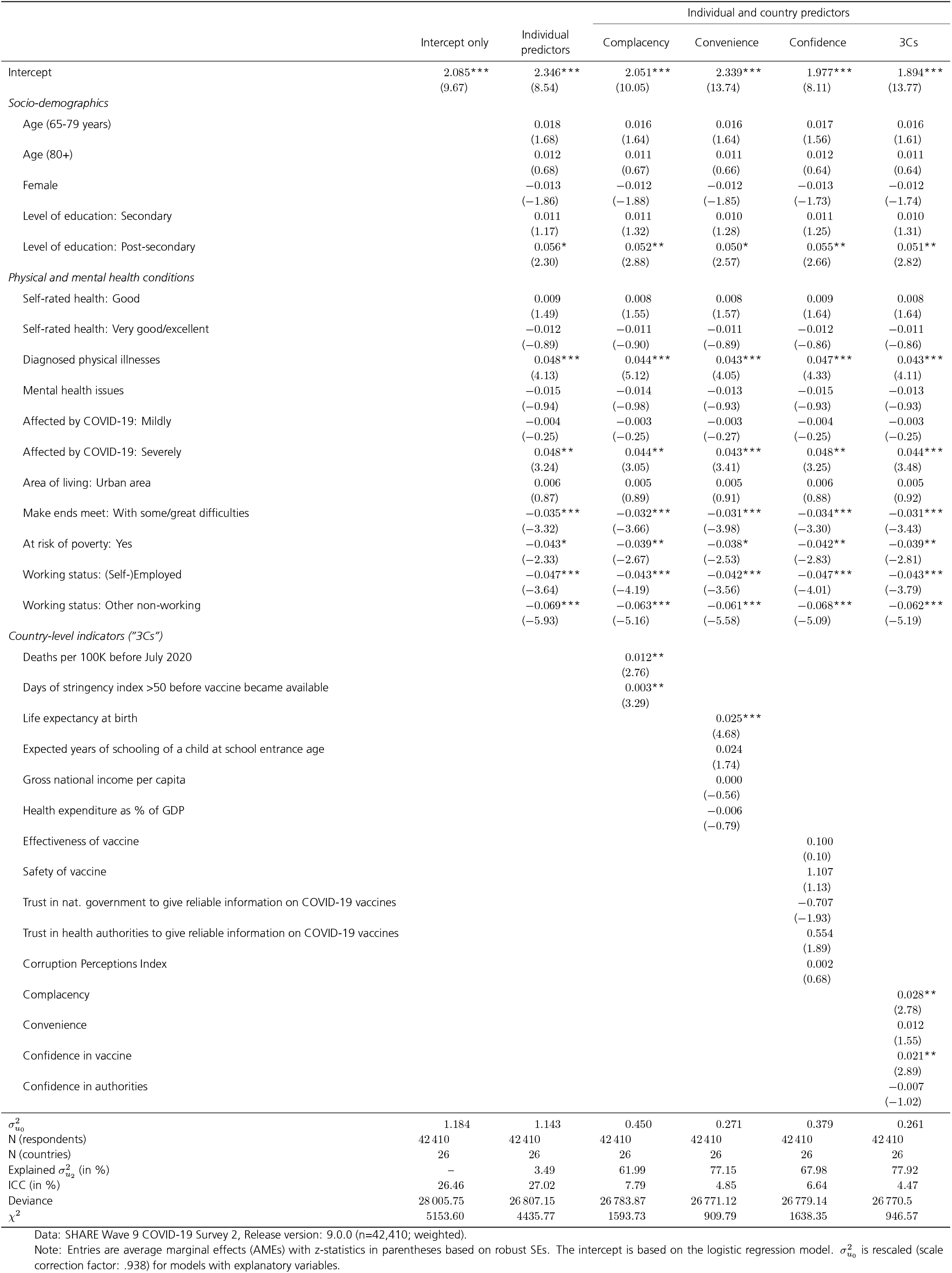
Multilevel (random intercept) logistic models for vaccination intention.

## Footnotes

1 https://hdr.undp.org/data-center/documentation-and-downloads

2 https://apps.who.int/nha/database/ViewData/Indicators/en

3 https://europa.eu/eurobarometer/surveys/detail/2512

4 https://www.transparency.org/en/cpi/2021

## Notes

### Competing Interest Statement

The authors have declared no competing interest.

### Author Declarations

The SHARE project and its data collection procedures are regularly reviewed prior to each new wave of data collection. The SHARE data collection procedures have been reviewed and approved by the Ethics Council of the Max Planck Society (MPG) every two years since 2012. The Ethics Council of the MPG has carefully reviewed the material of the SHARE project and confirmed that the overall research project and its procedures, the measures taken to ensure confidentiality and data protection, and the information provided to participants comply with international ethical standards. SHARE-ERIC's activities related to research involving human subjects are guided by international principles of research ethics such as the 'Respect Code of Practice for Socio-Economic Research' and the 'Declaration of Helsinki' (as prepared by the Council for International Organizations of Medical Sciences in collaboration with the World Health Organization, last revised at the 64th WMA Meeting in Fortaleza/Brazil in October 2013). In addition, the Max Planck Institute for Social Law and Social Policy has assured the Office for Human Research Protections (OHRP) of the U.S. Department of Health and Human Services (HHS) through its Federalwide Assurance (FWA) for the Protection of Human Subjects that all its activities involving research on human subjects are guided by the Declaration of Helsinki and the International Ethical Guidelines for Biomedical Research Involving Human Subjects.

## REFERENCES

Asparouhov, T. (2006). General Multi-Level Modeling with Sampling Weights. Communications in Statistics - Theory and Methods, 35(3), 439–460. 10.1080/03610920500476598

Bade, V., Schmitz, H., & Tawiah, B. B. (2024). Regional variations in vaccination against COVID-19 in Germany (S. Porcher, Ed.). PLOS ONE, 19(4), e0296976. 10.1371/journal.pone.0296976

Bergmann, M., Bethmann, A., Hannemann, T.-V., & Schumacher, A. T. (2022). Who are the Unvaccinated? Determinants of SARS-CoV-2 Vaccinations Among Older Adults Across Europe. easy_social_sciences, (Mixed 1), 1–11. 10.15464/easy.2022.01

Bergmann, M., & Wagner, M. (2021). The Impact of COVID-19 on Informal Caregiving and Care Receiving Across Europe During the First Phase of the Pandemic. Frontiers in Public Health, 9, 673874. 10.3389/fpubh.2021.673874

Bergmann, M., Wagner, M., Yilmaz, Y., Axt, K., Kronschnabl, J., Pettinicchi, Y., Schmidutz, D., Schuller, K., Stuck, S., & Börsch-Supan, A. (2024). SHARE Corona Surveys: Study profile. Longitudinal and Life Course Studies, 15(4), 506–525. 10.1332/17579597Y2024D000000027

Borga, L. G., Clark, A. E., D’Ambrosio, C., & Lepinteur, A. (2022). Characteristics associated with COVID-19 vaccine hesitancy. Scientific Reports, 12(1), 12435. 10.1038/s41598-022-16572-x

Börsch-Supan, A., Brandt, M., Hunkler, C., Kneip, T., Korbmacher, J., Malter, F., Schaan, B., Stuck, S., & Zuber, S. (2013). Data Resource Profile: The Survey of Health, Ageing and Retirement in Europe (SHARE). International Journal of Epidemiology, 42(4), 992– 1001. 10.1093/ije/dyt088

Carle, A. C. (2009). Fitting multilevel models in complex survey data with design weights: Recommendations. BMC Medical Research Methodology, 9(1), 49. 10.1186/1471-2288-9-49

Cipolletta, S., Andreghetti, G., & Mioni, G. (2022). Risk Perception towards COVID-19: A Systematic Review and Qualitative Synthesis. International Journal of Environmental Research and Public Health, 19(8), 4649. 10.3390/ijerph19084649

Franic, J. (2022). What Lies Behind Substantial Differences in COVID-19 Vaccination Rates Between EU Member States? Frontiers in Public Health, 10, 858265. 10.3389/fpubh.2022.858265

Goldstein, H., Browne, W., & Rasbash, J. (2002). Partitioning Variation in Multilevel Models. Understanding Statistics, 1(4), 223–231. 10.1207/S15328031US0104_02

Gomes, I. A., Soares, P., Rocha, J. V., Gama, A., Laires, P. A., Moniz, M., Pedro, A. R., Dias, S., Goes, A. R., Leite, A., & Nunes, C. (2022). Factors Associated with COVID-19 Vaccine Hesitancy after Implementation of a Mass Vaccination Campaign. Vaccines, 10(2), 281. 10.3390/vaccines10020281

Hale, T., Angrist, N., Goldszmidt, R., Kira, B., Petherick, A., Phillips, T., Webster, S., Cameron-Blake, E., Hallas, L., Majumdar, S., & Tatlow, H. (2021). A global panel database of pandemic policies (Oxford COVID-19 Government Response Tracker). Nature Human Behaviour, 5(4), 529–538. 10.1038/s41562-021-01079-8

Hox, J., Moerbeek, M., & Van De Schoot, R. (2010, September). Multilevel Analysis (0th ed.). Routledge. 10.4324/9780203852279

Jolly, N. A., Theodoropoulos, N., & Voucharas, G. (2025, March). Regional differences in COVID-19 vaccination rates across Europe. In K. A. Couch (Ed.), Handbook on Inequality and COVID-19 (pp. 52–71). Edward Elgar Publishing. 10.4337/9781035302765.00010

Lamot, M., & Kirbiš, A. (2024). Multilevel analysis of COVID-19 vaccination intention: The moderating role of economic and cultural country characteristics. European Journal of Public Health, 34(2), 380–386. 10.1093/eurpub/ckae022

Lunz Trujillo, K., Green, J., Safarpour, A., Lazer, D., Lin, J., & Motta, M. (2024). COVID-19 Spillover Effects onto General Vaccine Attitudes. Public Opinion Quarterly, 88(1), 97–122. 10.1093/poq/nfad059

Pronkina, E., Berniell, I., Fawaz, Y., Laferrère, A., & Mira, P. (2023). The COVID-19 curtain: Can past communist regimes explain the vaccination divide in Europe? Social Science & Medicine, 321, 115759. 10.1016/j.socscimed.2023.115759

Pronkina, E., & Rees, D. I. (2022, October). Predicting COVID-19 vaccination uptake (tech. rep. No. 15625). https://docs.iza.org/dp15625.pdf

Roghani, A. (2021). The relationship between macro-socioeconomics determinants and COVID-19 vaccine distribution. AIMS Public Health, 8(4), 665– 664. 10.3934/publichealth.2021052

Rughiniş, C., Vulpe, S.-N., Flaherty, M., & Vasile, S. (2022). Vaccination, life expectancy, and trust: Patterns of COVID-19 and measles vaccination rates around the world. Public Health, 210, 114–122. 10.1016/j.puhe.2022.06.027

Scherpenzeel, A., Axt, K., Bergmann, M., Douhou, S., Oepen, A., Sand, G., Schuller, K., Stuck, S., Wagner, M., & Börsch-Supan, A. (2020). Collecting survey data among the 50+ population during the COVID-19 outbreak: The Survey of Health, Ageing and Retirement in Europe (SHARE). Survey Research Methods, 217–221 Pages. 10.18148/SRM/2020.V14I2.7738

SHARE-ERIC. (2024a). Survey of Health, Ageing and Retirement in Europe (SHARE) Wave 1. Release version: 9.0.0. Data set. 10.6103/SHARE.w1.900

SHARE-ERIC. (2024b). Survey of Health, Ageing and Retirement in Europe (SHARE) Wave 2. Release version: 9.0.0. Data set. 10.6103/SHARE.w2.900

SHARE-ERIC. (2024c). Survey of Health, Ageing and Retirement in Europe (SHARE) Wave 3 - SHARELIFE. Release version: 9.0.0. Data set. 10.6103/SHARE.w3.900

SHARE-ERIC. (2024d). Survey of Health, Ageing and Retirement in Europe (SHARE) Wave 4. Release version: 9.0.0. Data set. 10.6103/SHARE.w4.900

SHARE-ERIC. (2024e). Survey of Health, Ageing and Retirement in Europe (SHARE) Wave 5. Release version: 9.0.0. Data set. 10.6103/SHARE.w5.900

SHARE-ERIC. (2024f). Survey of Health, Ageing and Retirement in Europe (SHARE) Wave 6. Release version: 9.0.0. Data set. 10.6103/SHARE.w6.900

SHARE-ERIC. (2024g). Survey of Health, Ageing and Retirement in Europe (SHARE) Wave 7. Release version: 9.0.0. Data set. 10.6103/SHARE.w7.900

SHARE-ERIC. (2024h). Survey of Health, Ageing and Retirement in Europe (SHARE) Wave 8. COVID-19 Survey Release version: 9.0.0. Data set. 10.6103/SHARE.w8ca.900

SHARE-ERIC. (2024i). Survey of Health, Ageing and Retirement in Europe (SHARE) Wave 8. Release version: 9.0.0. Data set. 10.6103/SHARE.w8.900

SHARE-ERIC. (2024j). Survey of Health, Ageing and Retirement in Europe (SHARE) Wave 9. COVID-19 Survey Release version: 9.0.0. Data set. 10.6103/SHARE.W9CA.900

Varbanova, V., Hens, N., & Beutels, P. (2024). Determinants of COVID-19 vaccination coverage in European and Organisation for Economic Co-operation and Development (OECD) countries. Frontiers in Public Health, 12, 1466858. 10.3389/fpubh.2024.1466858

WHO. (2014). Report of the SAGE Working Group on Vaccine Hesitancy (tech. rep.). World Health Organization. https://www.asset-scienceinsociety.eu/sites/default/files/sage_working_group_revised_report_vaccine_hesitancy.pdf

WHO. (2023). Vaccinating older adults against COVID-19 (tech. rep.). World Health Organization. https://iris.who.int/handle/10665/369450

WHO. (2024). WHO European Region, Emergency Operations Bulletin. First Quarter 2024 Weeks 1-13 (January-March 2024) (tech. rep.). World Health Organization. https://www.who.int/europe/publications/m/item/who-european-region-emergency-operations-bulletin-first-quarter-2024-weeks-1-13-january-march-2024

